# Delineating Drug Class and Target-Specific Adverse Events of Kinase Inhibitors

**DOI:** 10.1101/2024.08.07.24311573

**Authors:** Annalise Schweickart, Juhi Somani, Ryan Theisen, Navriti Sahni, Anna Cichońska, Rayees Rahman

## Abstract

Kinase inhibitors are a successful category of therapeutics used in treating diseases such as cancer, autoimmunity, and neurodegeneration. Despite their efficacy, these drugs often present clinically relevant adverse events that can limit their therapeutic utility or lead to their failure in clinical trials. The adverse event profile of a kinase inhibitor can be explained by its off- and on-target binding profile. Unfortunately, there are limited resources to couple inhibition of a specific kinase to clinical adverse events. Discerning which adverse events can be attributed to a specific kinase, and which are more generally caused by the kinase inhibitor drug class, is crucial for designing next-generation drugs that avoid toxicity and enhance clinical viability. By integrating adverse event incident data from the FDA Adverse Events Reporting Database with machine learning-predicted molecular binding profiles, we developed a statistical method that associates specific adverse events with potent inhibition of certain kinases. We also identify general adverse events inherent to the kinase inhibitor class. We validate our model through an extensive literature review of known kinase-adverse event pairs, comparison with the OnSIDES drug label side effect dataset, and prospective prediction of adverse events of recently approved kinase inhibitors. We show that our method can recapitulate well-established kinase-toxicity associations and identify previously unreported kinases associated with adverse events.

## INTRODUCTION

Protein kinase inhibitors (KIs) are a prominent class of targeted therapeutics primarily used in treating various diseases, especially cancer. To date, the US Food and Drug Administration (FDA) has approved 80 KIs for therapeutic use, and more than 180 are currently in clinical and preclinical development (*1*). Despite their clinical utility, many kinase drugs are associated with significant patient toxicity, such as QT prolongation (*2,3*), viral or bacterial infections (*4, 5*), and severe diarrhea (*6*). In clinical and preclinical stages, the observation of such serious adverse events (AEs) can limit the therapeutic index or impede the approval of such drugs. Consequently, the development of methods that can effectively model and mitigate the risk of specific AEs is vital for advancing the next generation of kinase therapeutics.

AEs associated with kinase drugs stem from a variety of causes, such as the chemical properties or toxophores of the inhibitors (*3,7*) or the inhibition of specific kinases (*8–11*). Many kinase drugs exhibit on-target toxicity due to potent engagement of a KI with its intended kinase targets (*4,8,12*). Conversely, due to the high degree of structural homology at the drug binding pocket (*13–15*), many AEs may result from unintended ‘off-target’ kinase inhibition by the KI (*16, 17*). While the design of kinase drugs typically strives for high target selectivity, achieving absolute specificity is challenging (*18–20*). In this context, polypharmacology—where a single drug interacts with multiple targets—may be not only an unavoidable outcome but also a requisite feature for efficacy (*21–23*). The complexity of kinase drug interactions underlines a main challenge in kinase drug design: achieving clinical efficacy while minimizing toxicity through precise target engagement.

Drug-associated AE prediction is a highly explored field of study, with popular approaches including analysis of chemical features, gene expression profiles, and network analysis (*24*). While these approaches leverage drug dose, pharmacokinetics, and genetics in their predictions, they do not address kinase target-specific association to AEs. Work by Gong et al. leveraged binding data against 442 kinases across 16 FDA approved KIs to connect kinase inhibition to specific AEs (*25*). Their analysis, although successful in validating some known associations, was limited by the small dataset and the lack of robust statistical methods to fully explore kinase-AE relationships. Similar limitations were observed in earlier work by Yang et al. where the authors calculated a kinase-AE association matrix from a dataset of only 20 KIs with more limited binding data, describing just 266 kinases (*26*). In a broader scope, the Secondary Pharmacology Database was developed to associate AEs with off-target interactions, testing 1,958 compounds across 200 safety pharmacology assays. However, among these 200 assays, only one specifically measured kinase inhibition (*27*). The lack of extensive binding data and statistical validation in these studies highlights a pivotal area for improvement, as both proper data and statistical frameworks are essential to strengthen the predictive power and scientific rigor of kinase-AE association models.

In this work, we extended the methods presented by Gong et al. and Yang et al. by deconvoluting kinase-AE associations from 112 KIs across 425 kinases (Fig. 1A). We integrated physician-reported AE data from the FDA Adverse Events Reporting System (FAERS) (*28*) and drug label AE data from the OnSIDES dataset (*29*) (Fig. 1A i). For each KI, we evaluated the overlap of AEs between FAERS and OnSIDES, and calculated the population reporting odds ratio for each AE in KIs compared to other drug target categories. We then applied a machine learning model to estimate the affinity of each KI across the human kinome. A kinase-AE association score was computed by integrating the KI-AE association matrix with the KI-kinase binding matrix. For each kinase-AE association score, we performed permutation testing to derive a representative p-value, evaluating the strength of the association (Fig. 1A ii). Finally, we evaluated our method by benchmarking kinase-AE associations both retrospectively via literature review, and prospectively using recently approved kinase drugs (Fig. 1A iii). Owing to the breadth and scope of the analysis, we anticipate this web-resource will support preclinical discovery and development of novel kinase therapeutics.

**Fig. 1.**
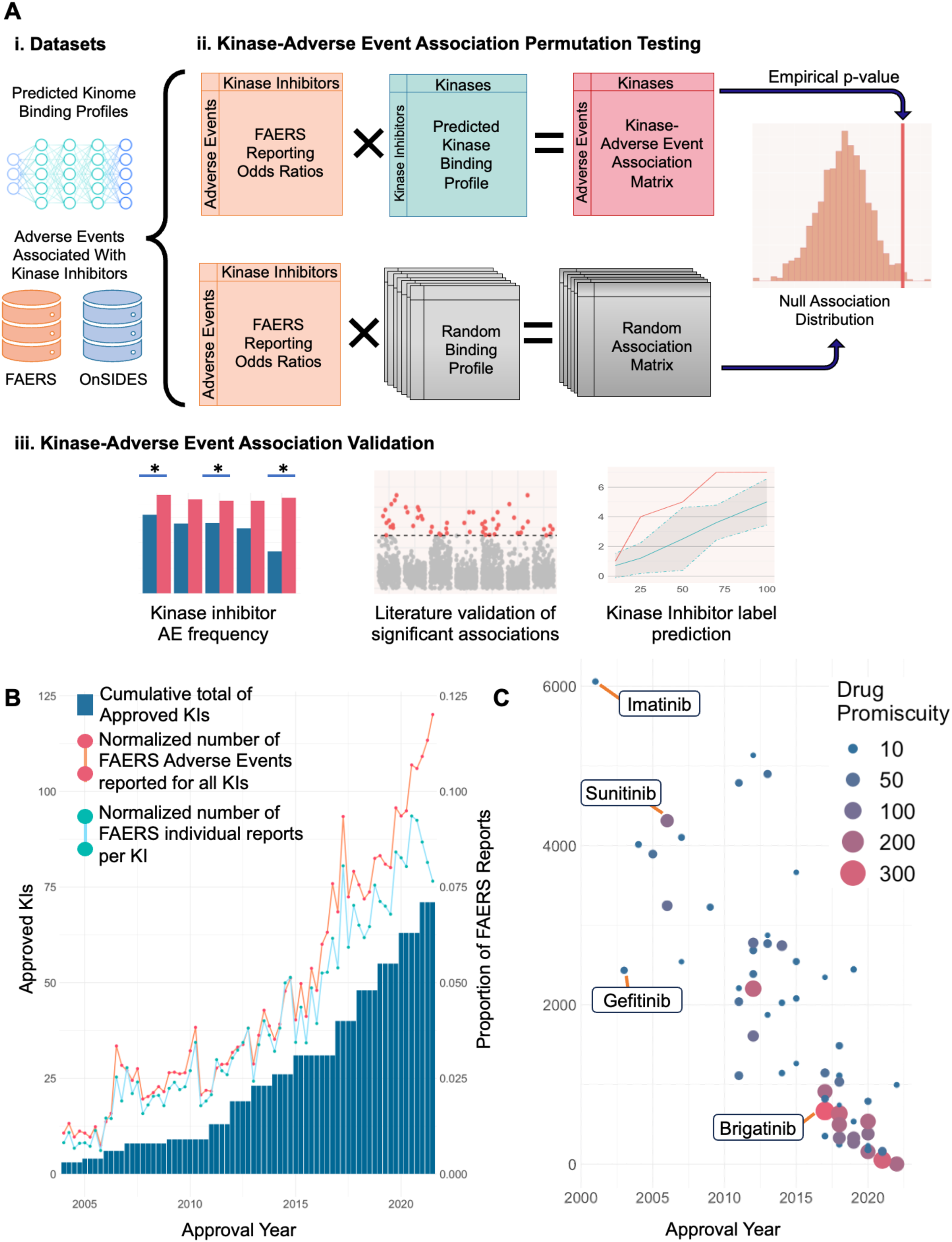
Overview of the Kinase-AE association method. (**A**) Predicted kinome binding profiles for 112 KIs are combined via matrix multiplication with the reporting odds ratios of those KIs for all AEs in the FAERS dataset. This results in an association score for each kinase-AE pair. This matrix multiplication is repeated 10,000 times on random kinome binding profiles to create a null association distribution against which the original score is compared to derive an empirical p-value. The AE space of KIs is explored by looking at AE frequencies within the FAERS dataset in KIs vs. non-KIs, comparing significant associations to known literature associations, predicting labels of KIs from significant associations and analyzing the chemical properties of KIs. (**B**) Cumulative total of approved KIs since 2000 (blue) accompanied by the non-cumulative number of AEs coinciding with KIs reported to the FAERS database normalized by all FAERS reports for the same time period (orange) and the normalized number of AE FAERS reports, per KI. (**C**) Number of unique AEs reported for each approved KI over time. Each point represents a KI whose size reflects the number of predicted kinase binders.

## RESULTS

### Adverse event reports for kinase inhibitor drugs are increasing, but not due to increased promiscuity

Since the 2001 approval of imatinib, over 80 KIs have been approved for a wide array of indications (*1*). From 2004 to 2021, as the number of approved KIs and their reported usage increased, there was a significant rise in both the absolute number of AEs reported for the KI therapeutic class as a whole and the average number of FAERS reports per KI in the FAERS database (Fig. 1B). This upward trend was further corroborated when we examined the emergence of new, previously unreported AE occurrences in KIs over time (Fig. S1A). Both the rate of previously unseen AEs and the overall rate of reported AEs for KIs have been increasing over time, compared to the stagnant rates observed in GPCR therapeutics (Fig. S1B).

Given the escalating trend in AEs linked to KIs, we aimed to determine whether this increase was due to the well-documented binding promiscuity of KIs. To investigate this, we utilized a custom machine learning model to predict the kinome-wide binding profile of all KIs in the FAERS dataset (see Methods) and compared promiscuity to unique AEs reported in FAERs within the first three years of approval for each KI. Surprisingly, we observed no correlation between the predicted promiscuity of KIs and the number of unique AEs observed (Fig. 1C). This result is further corroborated when controlling for the rate of new AEs observed per FAERS report, where each FAERS report can be considered as an individual KI prescription, and by the reported promiscuity of these molecules in literature (Fig S2). Relatively selective drugs, such as tofacitinib and ibrutinib, exhibited higher amounts of observed toxicity events compared to their more promiscuous counterparts, such as sunitinib and dasatinib. This lack of association between predicted promiscuity and AE frequency indicates that KI promiscuity may not be the primary driver of AEs.

### The AE space of kinase inhibitors shows key distinctions from other therapeutics

To identify AEs uniquely associated with the kinase inhibition, we conducted a comparative analysis. This involved evaluating the incidence rate of each AE in KIs, defined as the proportion of KIs with at least one AE report in FAERS, and comparing this rate to the incidence rate of the same AEs across all other drug categories (Fig. 2A, Tables S1-2). By contrasting these frequencies, we identified five rough categories of AEs.

**Fig. 2.**
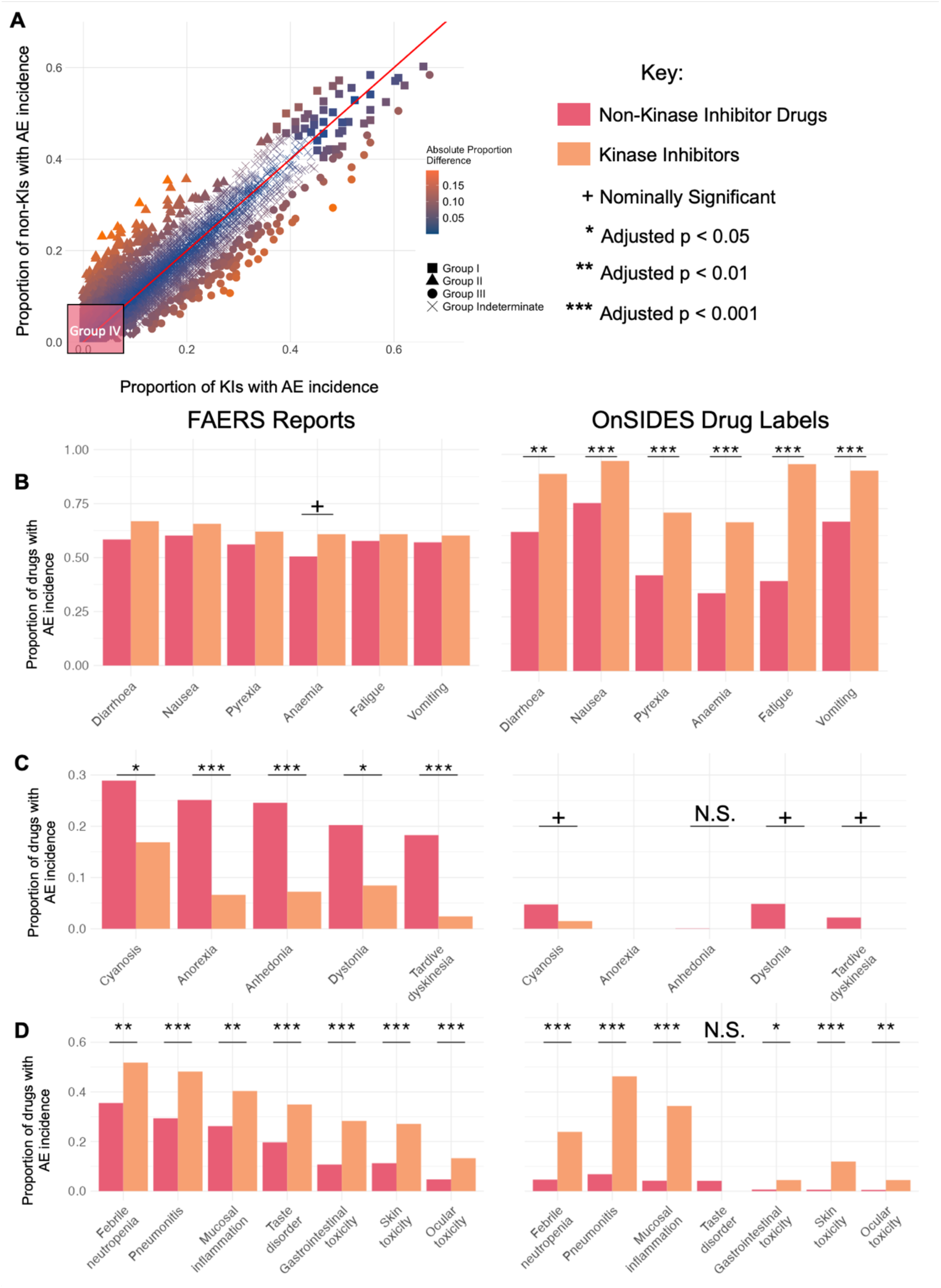
Delineating class-specific adverse events of kinase inhibitors. (**A**) Comparison of the proportion of KIs with a given FAERS AE with the proportion of non-KIs with that same event. Each point in this plot represents a single AE, with the color reflecting the difference between the two proportions. Each symbol represents an AE Group. (**B**) Proportions of selected drugs that coincide with Group I AEs, that is AEs that occur in over 45% of KIs in the FAERS dataset (KIs in orange, non-KIs in red). Proportions from the FAERs dataset are on the left, proportions from the OnSIDES drug labels dataset on the right. (**C**) Proportions of drugs that coincide with Group II AEs which occur at least 7.5% more frequently in non-KIs as compared to KIs in the FAERs dataset and their corresponding ratios in the OnSIDES dataset. (**D**) Proportions of drugs that coincide with Group III AEs that occur at least 7.5% more frequently in KIs as compared to non-KIs in the FAERs dataset and their corresponding ratios in the OnSIDES dataset. N.S. – Not Significant.

Group I AEs are events observed with >45% incidence in both KIs and non-KIs. Common AEs in this group include MedDRA (http://www.meddra.org/, refer to Methods) Preferred Terms (PTs), such as ‘diarrhea’, ‘nausea’, ‘pyrexia’, ‘anemia’, ‘fatigue’, and ‘vomiting’. Interestingly, despite having a similar reporting incidence in FAERS compared to other therapeutic categories, they were significantly more prevalent on drug labels of KIs than non-KIs (Fig. 2B). We propose that Group I AEs represent toxicities occurring at a generally high frequency, potentially stemming from kinase inhibition.

Group II AEs exhibit an incidence that is at least 7.5% higher in non-KIs compared to KIs (Fig. 2C). Events in this group include ‘cyanosis’, ‘anorexia’, ‘anhedonia’, ‘dystonia’, and ‘tardive dyskinesia’. Of particular prevalence are AEs associated with drug administration sites (Table S1). As most KIs are administered orally, they do not lead to administration-related AEs as frequently as non-KIs. These AEs are significantly enriched in non-KIs in both reporting incidence and label presence, suggesting they may not be directly attributable to any specific drug-kinase interaction.

In contrast, Group III are AEs that are at least 7.5% higher in incidence in KIs than non-KIs (Fig. 2D). Examples include ‘neutropenia’, ‘pneumonitis’, ‘gastrointestinal toxicity’, and ‘skin toxicity’, which are enriched in FAERS reporting frequency and label presence, indicating KI class specificity. For instance, neutropenia is an AE associated with on-target interactions with specific KIs like ruxolitinib and JAK2 inhibitors (*30*). Group III AEs may be closely linked to kinase inhibition within the kinome. While all FAERs reports in which the AE fell under the same MedDRA category as the treatment indication were omitted from this analysis (see Methods), AEs related to diseases commonly treated by KIs may still be found in Group III. This is evidenced by the large number of immune system and blood cell-related AEs, which could be related to the autoimmune disorders and heme malignancies KIs treat, respectively.

Finally, the remaining 17,000 primary MedDRA terms not represented by the previous groups were categorized into Group IV and Group Indeterminate. Group IV contains AEs that are infrequently reported in the FAERS database, each with an incidence rate of less than 10% for both KIs and non-KIs. Group Indeterminate comprises terms that do not show a differential occurrence between KI and non-KI groups (Fig. 2A). Collectively, these terms cover a wide range of concepts with varying degrees of relevance for modeling drug toxicity. In both categories, there exist terms with limited utility, such as ’physical fitness training’, ’bed rest’, ’bone marrow scan’, ‘apparent life-threatening event’, and ’bite.’ On the other hand, terms such as ’arthrotoxicity’, ’anoxic encephalopathy’, and ’injection site muscle atrophy’ are highly relevant for assessing drug toxicity. Without adequate representation in the FAERS dataset or a clear proportional bias towards KIs, categorizing these terms in relation to KI toxicity presents a significant challenge.

The data presented here suggests a nuanced landscape of AEs associated with KIs, distinguishing them from other drug categories (*31*). We hypothesize that our categorization of AEs may refine approaches that aim to deconvolute KI binding profiles and AE incidence, illuminating kinase anti-targets that drive observed toxicities.

### Curation of kinase-AE associations from literature

To learn the association of AEs with the inhibition of specific kinases, we constructed a comprehensive benchmark dataset through a manual review of 91 publications. Our extensive literature review resulted in 398 kinase-AE associations, as detailed in Table S3. Our dataset significantly surpasses Gong et al.’s catalog, which documented 27 kinase-AE associations from 16 references (*25*). While not exhaustive, our literature review offers valuable insights into the connection between kinase inhibition and toxicity.

Our analysis of this benchmark dataset reveals that a substantial proportion of AEs can be traced back to the inhibition of just a few kinases (Fig. 3A). For example, AEs such as epidermal and dermal conditions, gastrointestinal issues, cardiac arrhythmias, and anemias can be linked to just the VEGFR family of kinases (FLT1, KDR, FLT4, and FLT3) (Fig. 3A). Additionally, the reporting frequency of these AEs can vary, covering Groups I-IV (Fig. S3).

**Fig. 3.**
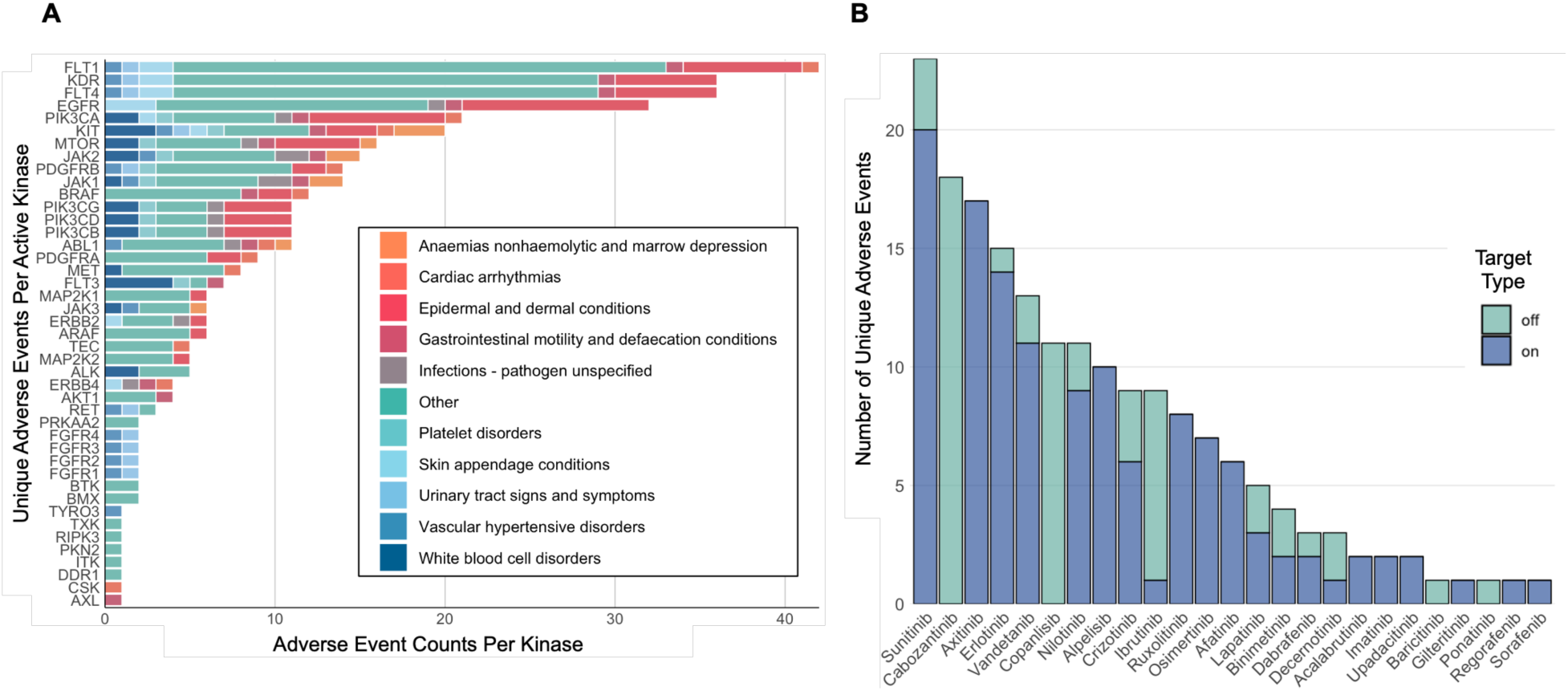
Analysis of KI-AE associations in literature. (**A**) Count of HLGT terms associated with each kinase in the benchmark kinase-AE set. B) Number of unique AEs observed in selected KIs attributable to either on-target or off-target interactions.

Since several toxicities are reported to be associated with a few kinases, we explored whether AEs linked to specific KIs can be attributed to their intended (on-target) or unintended (off-target) effects (Fig. 3B). Our findings suggest that many reported AEs can be related to on-target interactions. For example, sunitinib (Sutent), which has the highest number of reported AEs, is a VEGFR inhibitor, which, in turn, is associated with a wide range of AEs (Fig. 3A). This suggests that the toxicity of KIs often stems from their primary mechanisms of action rather than unintended interactions.

### Permutation testing recovers literature-curated kinase-AE associations and prospectively predicts AEs for newly approved drugs

To accurately associate kinases with toxic events, we developed a novel statistical approach. This involves calculating a kinase-AE association score based on the covariance between the relative odds ratio of an AE across all KIs in the FAERS database and the predicted binding affinity of a kinase for those KIs (see Methods). Through extensive permutation testing (Fig. 1A ii), we determined an empirical p-value for each kinase-AE association. We evaluated our method’s ability to recover literature-reported kinases associated with specific AEs using our benchmark dataset.

Our method demonstrated reasonable recall for benchmark kinases (Fig. 4A), successfully identifying all benchmark kinases within the top 10th percentile (by p-value of each kinase-AE association) for 24% of AEs (28 out of 116 PTs). Notably, at least half of all benchmark kinases were recovered in the top 10th percentile for 38% of benchmark AEs (44/116 PTs).

**Fig. 4.**
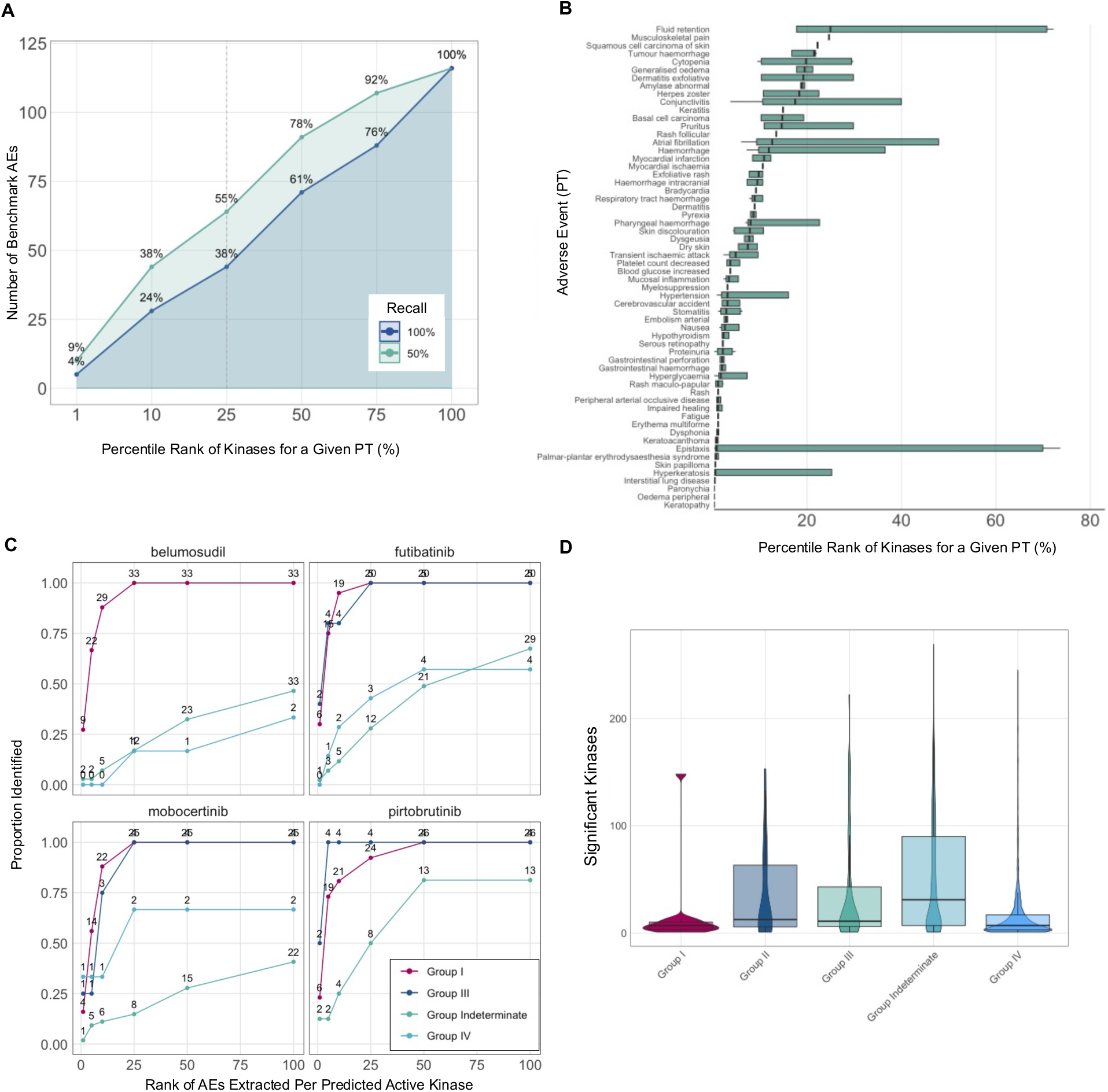
Retrospective and prospective evaluation of our method. (**A**) Number of benchmark AEs that have either 100% or 50% of literature associated kinases correctly identified as a function of the percentile rank of the kinase (as determined by our method). (**B**) Distributions of the percentile ranks of benchmark kinases plotted for each AE at the PT level of the MedDRA hierarchy. This is specifically for a subset of AEs where the median percentile rank is below 25%. (**C**) Predictive performance of identifying AEs present on labels for recently approved KIs not observed in our dataset: belumosudil, futibatinib, mobocertinib, pirtobrutinib across Group I,III, IV and Indeterminate AEs. (**D**) Total number of significant kinase associations per AE group.

Our method excelled in identifying kinases linked to AEs such as hyperkeratinosis and peripheral arterial occlusive disease (Fig. 4B) but was less effective for AEs like photosensitivity reactions and infections (Fig. S4). Evaluation at the highest MedDRA classification level, HLGT, revealed strengths in detecting tissue disorders and respiratory symptoms but showed limitations in associating kinases with immune and vision disorders (Fig. S5). This variability in predictive accuracy across different MedDRA categories underscores the challenge of analyzing kinase-mediated toxicities within existing classification systems, potentially hindering deeper analysis into these toxicities.

Having demonstrated the effectiveness of our kinase-AE association method retrospectively using our benchmark kinase dataset, we aimed to assess its ability to prospectively predict AEs for recently approved KIs (Fig. 4C). We selected four recently FDA-approved KIs for evaluation, each with a distinct mechanism of action and not included in the training of our method:

1. Belumosudil, a ROCK2 inhibitor, approved in 2021(*32*).
2. Mobocertinib, an EGFR inhibitor, approved in 2021(*33*).
3. Futibatinib, a FGFR2 inhibitor, approved in 2022(*34*).
4. Pirtobrutinib, a BTK inhibitor, approved in 2023(*35*).

For each KI, we predicted its kinome-wide binding profile using our kernel ridge regression model (Methods). We selected kinases deemed active by our method (predicted IC50 <1000 nM) and confirmed that our method accurately predicted the activity of each KI’s known primary target. We ranked the MedDRA PTs associated with the predicted active kinases by p-values and selected the top 1, 5, 10, 25, 50, and 100 AEs for each. We then collected the PTs listed on each KI’s label using the OnSIDES database.

Our method successfully identified 75-100% of Group I and III AEs within the top five ranked AEs per kinase (Fig. 4C) but performed poorly with Group Indeterminate AEs, as expected due to their less clear kinase-AE associations. Notably, it showed strong performance in identifying rare Group IV AEs for futibatinib and mobocertinib. This variability can be explained by the number of significant kinase associations for each AE group (Fig. 4D); Group I, III, and IV AEs, having fewer kinase associations on average, are more directly linked to specific kinase inhibition, and thus easier to predict. In contrast, Group II and Group Indeterminate AEs often have less clear or more complex associations with multiple kinases, increasing the challenge in accurately identifying these AEs.

Finally, we reviewed the top-ranked AEs for specific inhibitors and identified their associated kinase. For mobocertinib, dermatitis acneiform, mucosal inflammation, pneumonitis, and nail bed tenderness ranked highest. These AEs were predicted to be linked to on-target EGFR inhibition (dermatitis acneiform, mucosal inflammation, nail bed tenderness) and off-target ALK inhibition (pneumonitis). Importantly, ALK inhibition is observed with osimertinib, a close analog of mobocertib (*33, 36*). Interestingly, nail AEs were also prevalent at top ranks for futibatinib, but were associated with predicted CSNK1A1 inhibition, which futibatinib has been indeed recently reported to inhibit >50% at a 100nM concentration (*34*). In both cases, predicted off-target interactions contributed to these toxicities.

Taken together, these results demonstrate that our kinase-AE association method can retrospectively identify known kinase-AE associations and prospectively predict AEs in newly approved KIs. It showed high accuracy in identifying both frequent and rare AEs, highlighting the potential of our approach for improving drug safety monitoring.

### Predicted kinase-AE associations show shared kinase family behavior

Having validated our approach, we reviewed the top kinase-AE associations generated by our method. We found that certain AEs had multiple significant kinase associations (FDR-adjusted p < 0.2) at all MedDRA hierarchy levels (Table S4-6). When examining AEs at the HLGT level, many of them showed an interesting pattern of behavior where kinases within the same family associated strongly with the same events (Fig. 5A), an observation mirrored by our benchmark kinase analysis (Fig. 3A). For example, while inhibition of EGFR has been implicated in causing skin fissures and corneal epithelial lesions (*37, 38*), our analysis showed elevated associations of EGFR as well as other family members, including ErbB2 and ErbB4, with these AEs (Fig. 5A i). This family-based pattern of association persisted for the RAF, PI3K, and EPH kinase families (Fig. 5A ii-iv). For instance, our method recovered the association between EPH and vasomotor conditions, supported by evidence that the EPH family is functionally linked to vascular morphogenesis and angiogenesis (Fig. 5A iv) (*39*). This was found despite only a few approved EPH-targeting KIs.

**Fig. 5.**
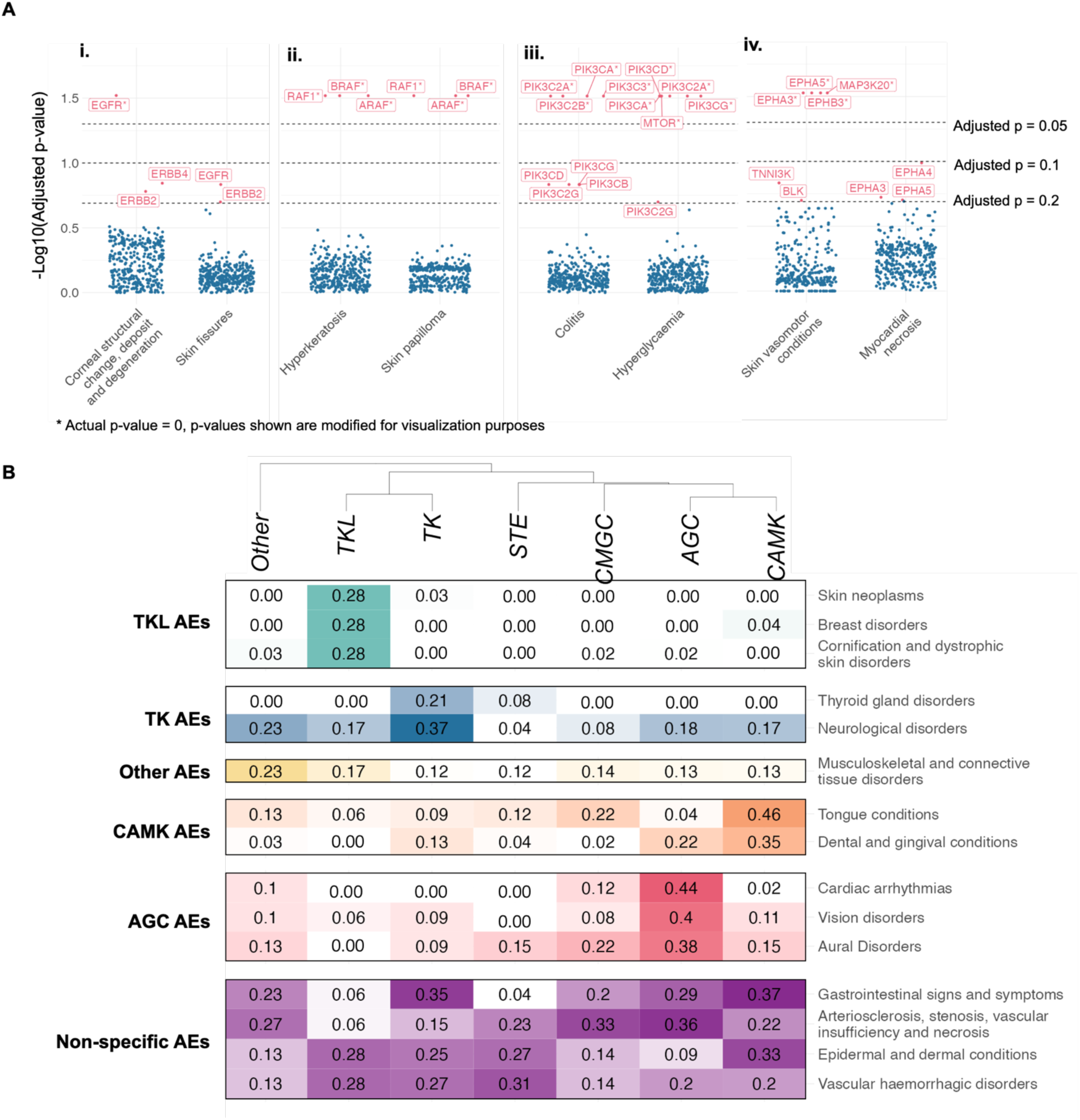
Kinase family and group-based associations to AEs. (**A**) Top kinases associated with select HLGT AEs that show shared family behavior. i. AEs associated with the Epidermal Growth Factor Receptor (EGFR) family, ii. AEs associated with the Rapidly Accelerated Fibrosarcoma (RAF) family, iii. AEs associated with the Phosphoinositide 3-kinase (PI3K) family, and iv. AEs associated with the Ephrin (EPH) family. (**B**) Clustering of kinase protein groups that are enriched with kinases associated with AEs at an HLGT level. Numbers inside cells represent the proportion of kinases within each protein group that show association (p < 0.2) with the HLGT.

To examine how broadly this family behavior extends, we zoomed out to the protein kinase group level to look for AEs common within homologous kinases (Fig. 5B). We observed the enrichment of certain specific AEs within the same kinase group. Tyrosine Kinase-Like (TKL) protein kinases are more enriched with kinases associated with skin neoplasms, breast disorders, and cornification and dystrophic skin disorders as compared to any other protein group, while Tyrosine Kinase proteins are more enriched for kinases associated with neurological disorders than any other protein group.

From this group-based analysis, we uncovered that certain AEs at the HLGT level are enriched across multiple protein kinase groups (Fig. 5B) –specifically epidermal and dermal conditions (“Skin toxicity”) and gastrointestinal signs and symptoms (“Gastrointestinal toxicity”). This could be because KIs with vastly different binding profiles across protein families can still trigger similar side effects. This is demonstrated by the associations of different kinase families to distinct types of skin rashes. For example, inhibition of EGFR was highly associated with skin fissures, RAF with hyperkeratosis and skin papilloma, and EPH with skin vasomotor conditions (Fig. 5A i, ii, iv). Although these varied skin conditions are often grouped under the general term “Rash” on drug labels—a term found on over 91% of KI labels in the OnSIDES dataset—our study reveals that the term “Rash” encompasses a range of distinct mechanisms at the kinase level, thus demonstrating the need for higher specificity in kinase AE labeling.

## DISCUSSION

Our work dives into the complexities of the AE space associated with protein KIs, aiming to inform more rational drug design strategies by identifying specific kinase-AE associations.

We observed that KIs are associated with an increasing number and variety of AEs over time compared to other drug classes. Intriguingly, this increase in AEs within the KI class does not appear to be related to the overall promiscuity of the inhibitors, therefore challenging the conventional perspective on drug off-target interaction landscape and its subsequent AEs.

Our analysis differentiates itself from preceding studies by categorizing AEs into several classes relevant in the context of kinase-AE prediction. We identified AEs that occur across a majority of KIs, which may be attributed to the inhibition of any given kinase. For these events, existing literature demonstrates a causative link between kinase activity and the observed toxicity. An example of this is the association of skin and gastrointestinal toxicity with VEGFR inhibition. Notably, our analysis also delineates a category of events that should not be considered in this type of AE landscape, such as toxicities related to the administration of the drug, due to their disproportionate occurrence in non-KIs as compared to KIs.

To evaluate the statistical significance of our kinase-AE associations, we conducted rigorous permutation testing. The utility of our approach is underscored by its strong performance in recapitulating literature-derived kinases associated with several types of AEs, as well as prospective prediction of Groups I, III and IV AEs in the labels of recently approved drugs outside of our dataset. Certain AE groups, such as mucosal inflammation and hyperkeratinosis, were more accurately associated with KI activity, providing insights that have significant translational implications. Importantly, by deriving empirical p-values for each kinase-AE association, our approach extends previous research in the field, which has previously associated AEs with kinases using non-statistical metrics, such as information gain and feature importance from random forest machine learning models.

Further granularity is provided through our discovery and examination of kinase family and group-based patterns. This analysis allows us to uncover distinct molecular mechanisms that contribute to the occurrence of common AEs across different kinase families, such as the EGFR, RAF, PI3K, and EPH families. On the other hand, it is crucial to note that certain toxicities appear to be specific to particular kinase families, such as the JAK family’s differential effects on immune system related AEs (*12*). Such family behavior could have two possible explanations: first, kinases from the same family may colocalize in tissue to perform similar functions, and thus their inhibition may lead to similar dysfunctions. Second, inhibition of a specific kinase within a family is directly responsible for an AE, but structural similarities of family members lead to co-inhibition by KI drugs. Recent conformational modeling of protein kinase binding pockets has shown strong similarity between families, supporting the latter explanation (*40*).

Our study is not without limitations. First, we do not capture the full diversity of the kinome; our focus is primarily on the protein kinome, leaving out some off-target interactions with lipid and carbohydrate kinases, among others, that are not well characterized. Secondly, our KI activity prediction model serves as an approximation and cannot substitute for comprehensive, kinome-wide experimental data. Next, many AEs reported in FAERS and OnSiDES for KIs could be attributed to underlying disease symptoms rather than drug-induced toxicity, potentially confounding our analysis. While delineating these types of toxicities is outside the scope of this work, it may further refine the space of AEs relevant for KI toxicity modeling. Furthermore, utilizing FAERS data introduces certain biases into our findings, as the number of catalogued AEs highly correlates with the time duration for which a drug has been approved. Lastly, there is significant room for improving the terms used for classifying AEs to KIs. Our current implementation leverages the MedDRA classification system for AEs. This system presents complexities that may obfuscate our statistical approach, such as the one-to-many mapping of primary terms to higher lever group terms. We anticipate the use of more advanced natural language processing techniques that could learn functional relationships between events could potentially lead to a development of a more optimized system for classifying AEs.

In conclusion, our study illuminates critical aspects of the AE landscape for KIs, thus providing vital data for more rational and targeted drug design. Our findings present a nuanced understanding of kinase-associated AEs, which could contribute to the optimization of therapeutic strategies and mitigate adverse impacts, ultimately benefiting patient outcomes.

## MATERIALS AND METHODS

### Datasets

Food and Drug Administration Adverse Event Reporting System: The Food and Drug Administration Adverse Event Reporting System (FAERS) database is one of the largest pharmacovigilant databases in the world, cataloguing hundreds of millions of spontaneous AE reports submitted to the FDA by healthcare professionals, consumers, and pharmaceutical companies (*28*). Each AE catalogued is accompanied by the patient’s drug regimen, disease indication, and the date of report. For this analysis, over 170 million AE reports from the first quarter of 2004 through the third quarter of 2021 were extracted and processed. From FAERS, we extracted 6,777 small molecule AE profiles, among which 168 were KIs.

Medical Dictionary for Regulatory Activities: The Medical Dictionary for Regulatory Activities (MedDRA, http://www.meddra.org/) system version 25.1 was utilized to classify and analyze AEs in this study. MedDRA is a standardized hierarchical medical terminology developed specifically for the classification and coding of AEs. The MedDRA hierarchy consists of five levels: System Organ Class (SOC), High-Level Group Term (HLGT), High-Level Term (HLT), Preferred Term (PT), and Lowest-Level Term (LLT).

The Observational Safety in Data from Electronic Health Records dataset: The Observational Safety in Data from Electronic Health Records (OnSIDES) dataset was used in this study for independent validation. Labels were taken from the DailyMed FDA label database and preprocessed as previously described (*29*).

### Data Filtration

To ensure the inclusion of only drug-induced AEs in our analysis, we applied several filtering steps. Firstly, AEs that were likely caused by the underlying indication rather than the drug itself were removed. This was determined by comparing the indication of the FAERS report and AE classifications under the same High-Level Group Term (HLGT) umbrella in the MedDRA hierarchy. If the indication and AE fell within the same HLGT, indicating a potential association between the disease and the AE, those events were excluded.

Secondly, AEs falling under the System Organ Class (SOC) annotation of “Congenital, familial and genetic disorders” were eliminated from the analysis. These events were presumed to be genetically based rather than drug-induced and, therefore, were deemed not relevant for assessing drug safety. Additionally, AEs falling under the SOC annotation of “Social circumstances” were removed from the dataset. These events, such as the death of a family member or job loss, were considered unrelated to drug use and were not considered in the analysis.

Following these filtration steps, the dataset was refined to include 145,037,262 drug-AE pairs that were likely attributable to drug use. These filtered pairs were used for subsequent analyses, enabling a focused investigation of drug-related AEs.

### Reporting Odds Ratio and AE Frequency Calculation

The Reporting Odds Ratio (ROR) was calculated for each of the remaining drug-AE pairs to identify potential disproportional associations between specific drugs and AEs. For each drug and AE the following calculation was performed at the PT, HLT, and HLGT MedDRA levels:

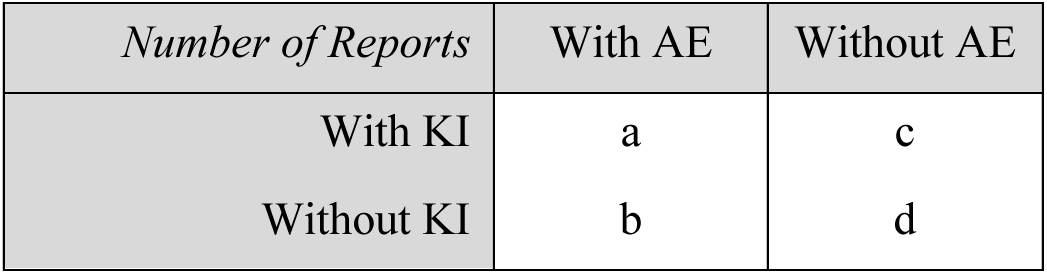

| *Number of Reports* | With AE | Without AE |
| --- | --- | --- |
| With KI | a | c |
| Without KI | b | d |

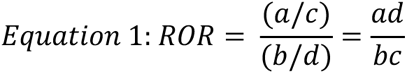

Resulting ROR scores were then log-transformed before subsequent analyses.

We identified a strong bias in our ROR calculations related to time of drug approval; newer drugs have less time to accumulate AE reports, decreasing term c in the ROR calculation. To counteract this bias, log-transformed ROR terms were then linearly modelled against time (Fig. 1 C,D), and the time-independent residuals from this model were used for all following analyses.

### Kinase Inhibitor Kinome Binding Prediction

To predict the kinome-wide binding affinity of an input small molecule. we leveraged a custom pairwise kernel ridge regression machine learning model, similar to the models described elsewhere (*41, 42*). We trained the model on compound-kinase Kd, Ki and IC50 data from ChEMBL32 (*43*) and PubChem (*44*), together with ECFP4 fingerprints and amino acid sequences of protein kinase ATP binding pockets. We evaluated the performance of the model on the hold-out set under two prediction scenarios applicable in our work: (i) filling in the gaps in otherwise known compound-kinase binding affinity matrix (“Compound-Kinase Spilt”), and (ii) predicting kinome-wide binding of a new compound scaffold not seen in the model training (“Compound Cluster Spilt”). Pairwise kernel ridge regression model demonstrates best-in-class performance compared to other compound-kinase prediction models benchmarked specifically, BiMCA (*45*), DeepDTA (*46*) and random forest (*47*) (Table 1). Importantly, for each point estimate of compound-kinase affinity, we leverage the connection between kernel ridge regression and Gaussian process to calculate an associated variance estimate which serves as a metric of the model’s uncertainty (*48*):

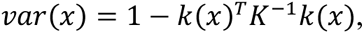

**Table 1.**
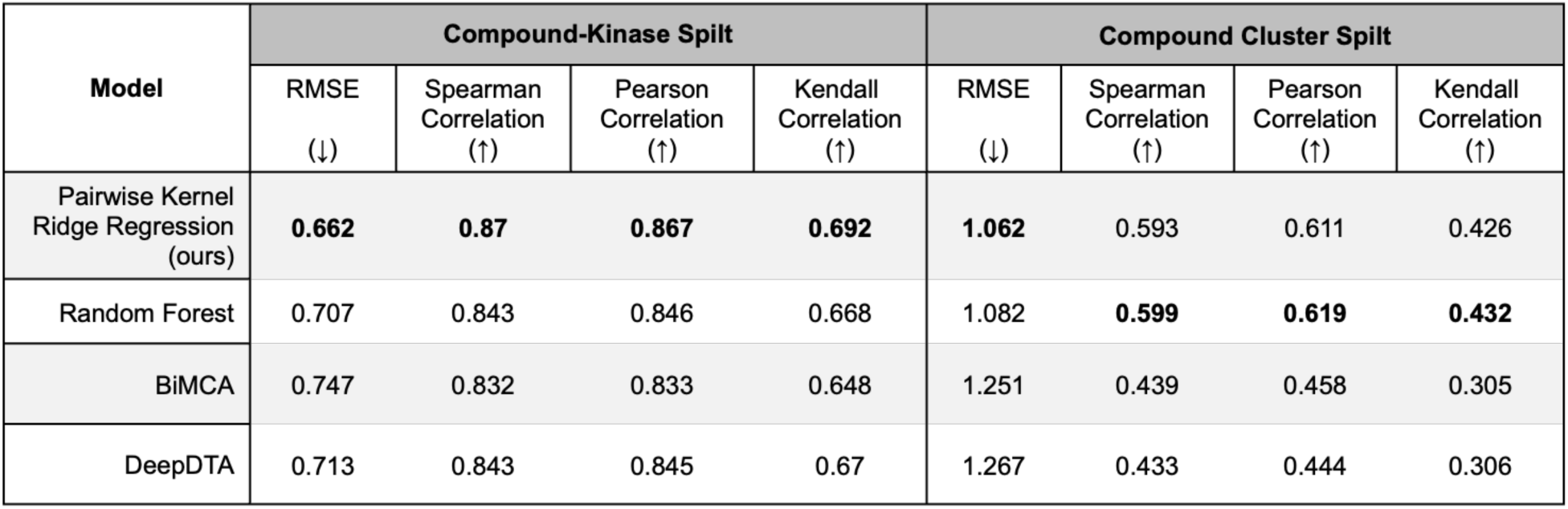
Performance of machine learning-based models to predict kinome-wide drug activity under distinct validation splits. Our approach, Pairwise Kernel Ridge Regression outperforms Random Forest, as well as published methods such as BiMCA and DeepDTA for predicting unseen compound-kinase pairs (Compound-Kinase Split) and performs favorably predicting activities for previously unseen chemotypes (Ligand Cluster Split). More information about this method can be found in Theisen et al. 2024

| Model | Compound-Kinase Split |  |  |  | Compound Cluster Split |  |  |  |
| --- | --- | --- | --- | --- | --- | --- | --- | --- |
|  | RMSE<br>(↓) | Spearman<br>Correlation<br>(↑) | Pearson<br>Correlation<br>(↑) | Kendall<br>Correlation<br>(↑) | RMSE<br>(↓) | Spearman<br>Correlation<br>(↑) | Pearson<br>Correlation<br>(↑) | Kendall<br>Correlation<br>(↑) |
| Pairwise Kernel Ridge Regression (ours) | **0.662** | **0.87** | **0.867** | **0.692** | **1.062** | 0.593 | 0.611 | 0.426 |
| Random Forest | 0.707 | 0.843 | 0.846 | 0.668 | 1.082 | **0.599** | **0.619** | **0.432** |
| BiMCA | 0.747 | 0.832 | 0.833 | 0.648 | 1.251 | 0.439 | 0.458 | 0.305 |
| DeepDTA | 0.713 | 0.843 | 0.845 | 0.67 | 1.267 | 0.433 | 0.444 | 0.306 |

where x = (c,p) is a new compound-protein pair, K is the training kernel, and k(x) = k(x, Xtrain) is the new pair-training sample kernel. We delineate likely predicted compound-kinase interactions, in predicted negative log activity (such as Kd, Ki, IC50) units (pActivity) from unlikely predicted interactions by thresholding based on the variance estimate and predicted pActivity (pActivity ≥ 6 and variance ≤ 0.7) (*49*).

### Adverse Event Classification and Association with Kinases

To find AEs specific to the KI drug class, we quantified the frequency of AEs within the FAERS and OnSIDES datasets associated with KI drugs, as defined by the Protein Kinase Inhibitor Database (PKIDB) versus non-KI small molecule drugs (*50*, *51*). Briefly, for each AE at the PT, HLT, and HLGT MedDRA levels, we count the number of KIs that have a catalogued co-incidence with that event. This number is then divided by the total number of KIs in the FAERS dataset. The same process is performed on non-KI drugs, creating a KI class-based proportion and a general drug proportion for each AE. This process is subsequently performed on the OnSIDES dataset in the same manner. Class-specific AEs were then determined by comparing the proportions between KI and non-KI drugs.

#### Kinase-specific Adverse Events

To find AEs associated with the inhibition of a specific kinase, we combined the kinome binding profiles of KIs in FAERS with their RORs to find covariance between kinases and AEs. A non-parametric permutation test was used to determine the underlying distribution of these covariances, against which we compared the observed covariance for each kinase-AE pair. This was performed as follows: First, a FAERS ROR matrix was created by converting the log-transformed ROR scores from the FAERS dataset to an AE by KI matrix. Next, a predicted kinase profile matrix was created by converting the KI binding profiles to a KI by kinase matrix. These two matrices were multiplied, resulting in an AE by kinase matrix containing a score for each AE-kinase pair (Fig. 1B). To find the underlying distribution of each of these scores, the FAERS ROR matrix was multiplied by a randomized kinase binding profile matrix where the binding values for each kinase were randomized. This kept the kinase-specific binding distributions the same, but each drug was given a shuffled binding profile. The result of this multiplication was a random association matrix. This randomization step was performed 5,000 times, resulting in a null association distribution for each kinase-AE pair. Finally, the real kinase-AE score was compared against this random distribution to calculate an empirical p-value, which was then FDR adjusted (Fig. 1C).

## Data Availability

All data produced in the present work are contained in the manuscript

## List of Supplementary Materials

Fig. S1 to Fig S4 (PDF)

Tables S1 to S6 (Excel file)

## Acknowledgments

We would like to thank our scientific advisors, Drs. Olivier Elemento and Andreas Bender for their input and feedback into the manuscript.

## Funding

None

## Author contributions

Conceptualization: AS, AC, RR Methodology: AS, JS, RT, AC, RR Investigation: AS, JS, RT, NS, AC, RR Visualization: AS, JS, RT, AC, RR Funding acquisition: RR Project administration: AC, RR Supervision: AC, RR Writing – original draft: AS, AC, RR Writing – review & editing: AS, AC, RR

## Competing interests

All authors were employees at Harmonic Discovery Inc. during the course of the study.

## Data and materials availability

All code used to generate figures and data will be available at https://github.com/Harmonic-Discovery.

## Notes

### Funding Statement

This study did not receive any funding

