## Supplementary Figures for "Delineating Drug Class and Target-Specific Adverse Events of Kinase Inhibitors"

Annalise Schweickart *et al.*

**This PDF file includes**

Figs. S1 to S4

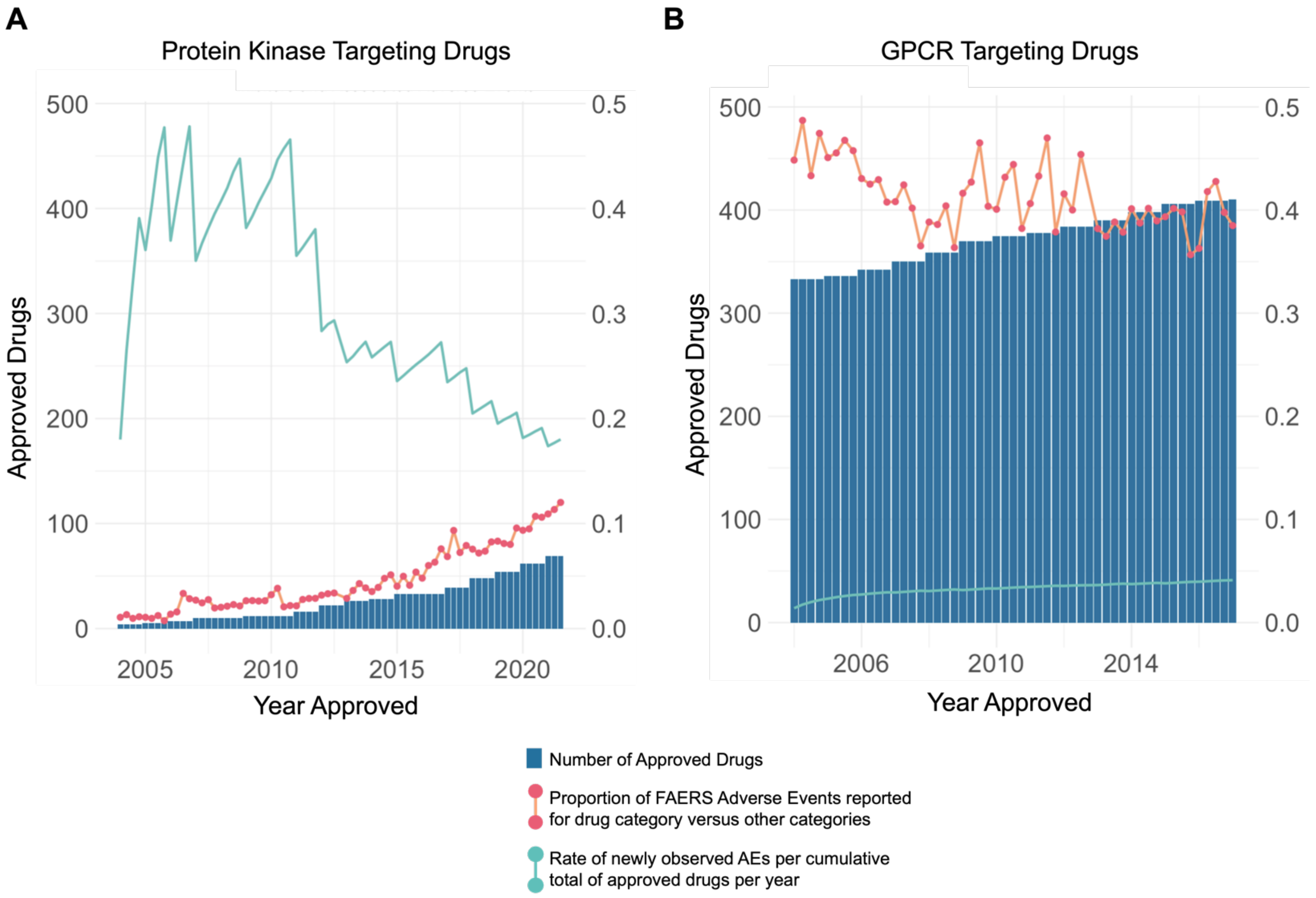


**Figure S1.** Trends in AE observation in KIs (A) and GPCR therapeutics (B) over time.

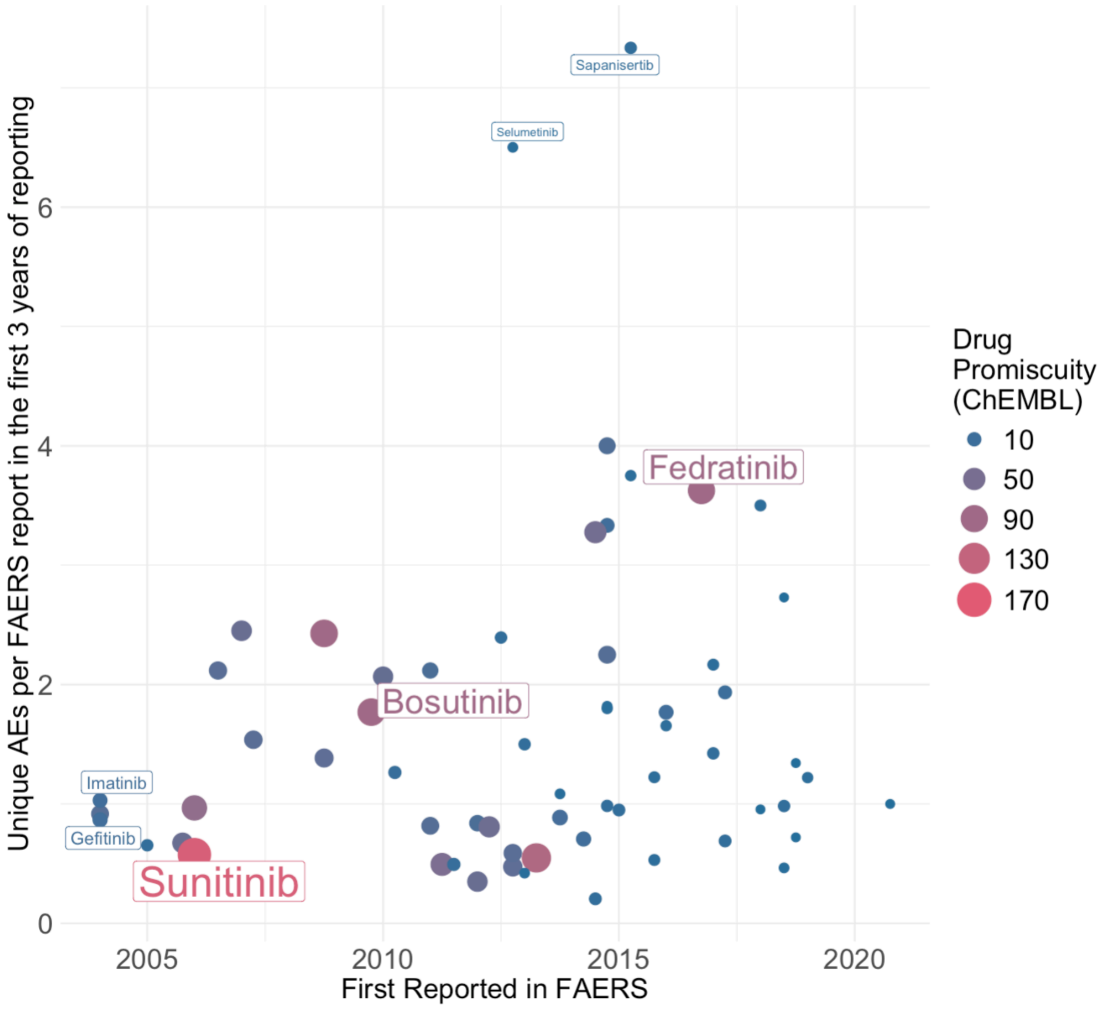


**Figure S2.** Rate of new AE observations, per FAERS report, within the first three years of a KI approval against the first approval date of the drug. The observed KI promiscuity, as determined by the number of ChEMBL reported kinase binders (IC50 <1,000nM) is reflected by the size and color of each point.

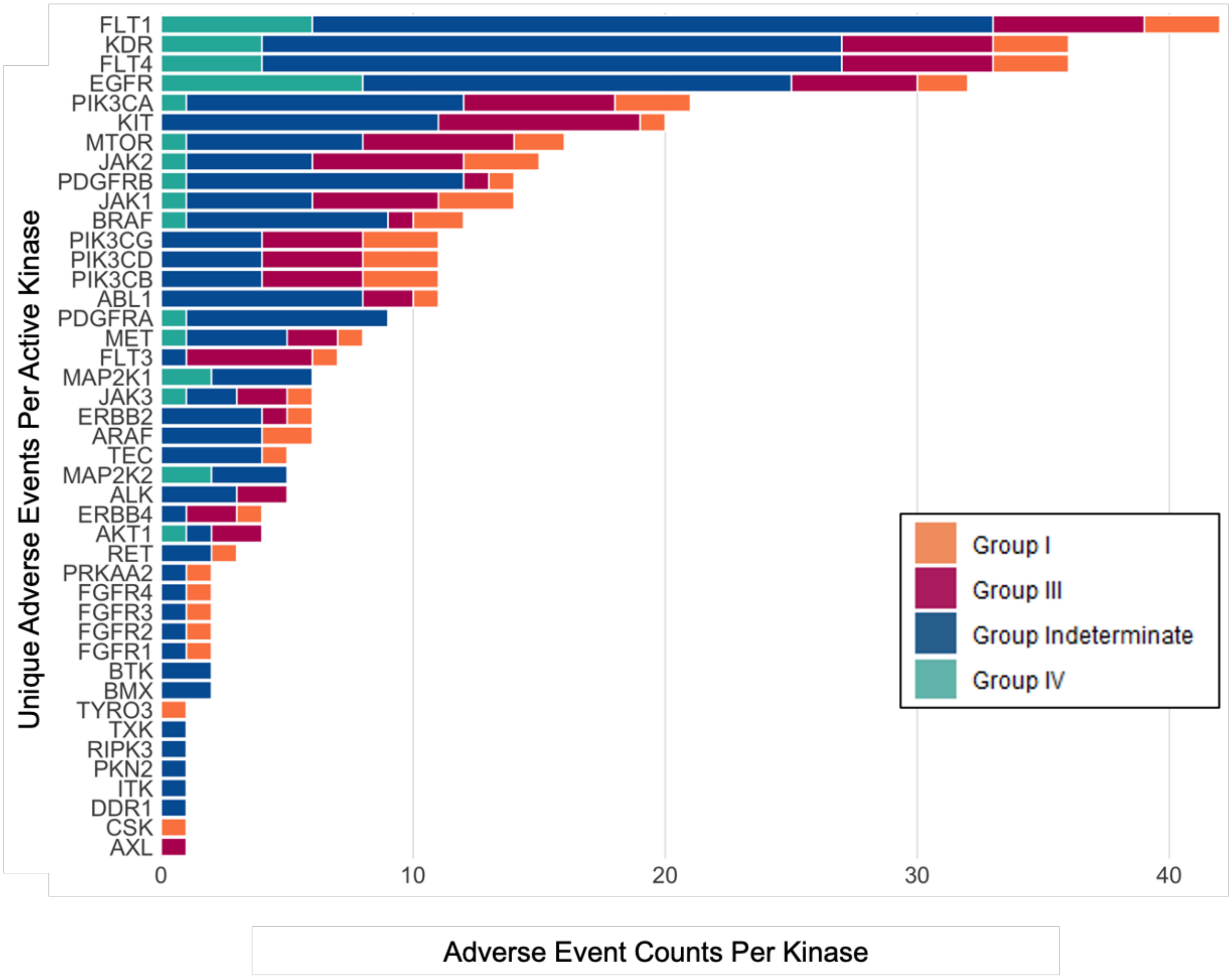


**Figure S3.** Count of AEs in each group, as delineated in Fig. 2A, for each kinase in the benchmark kinase-AE dataset.

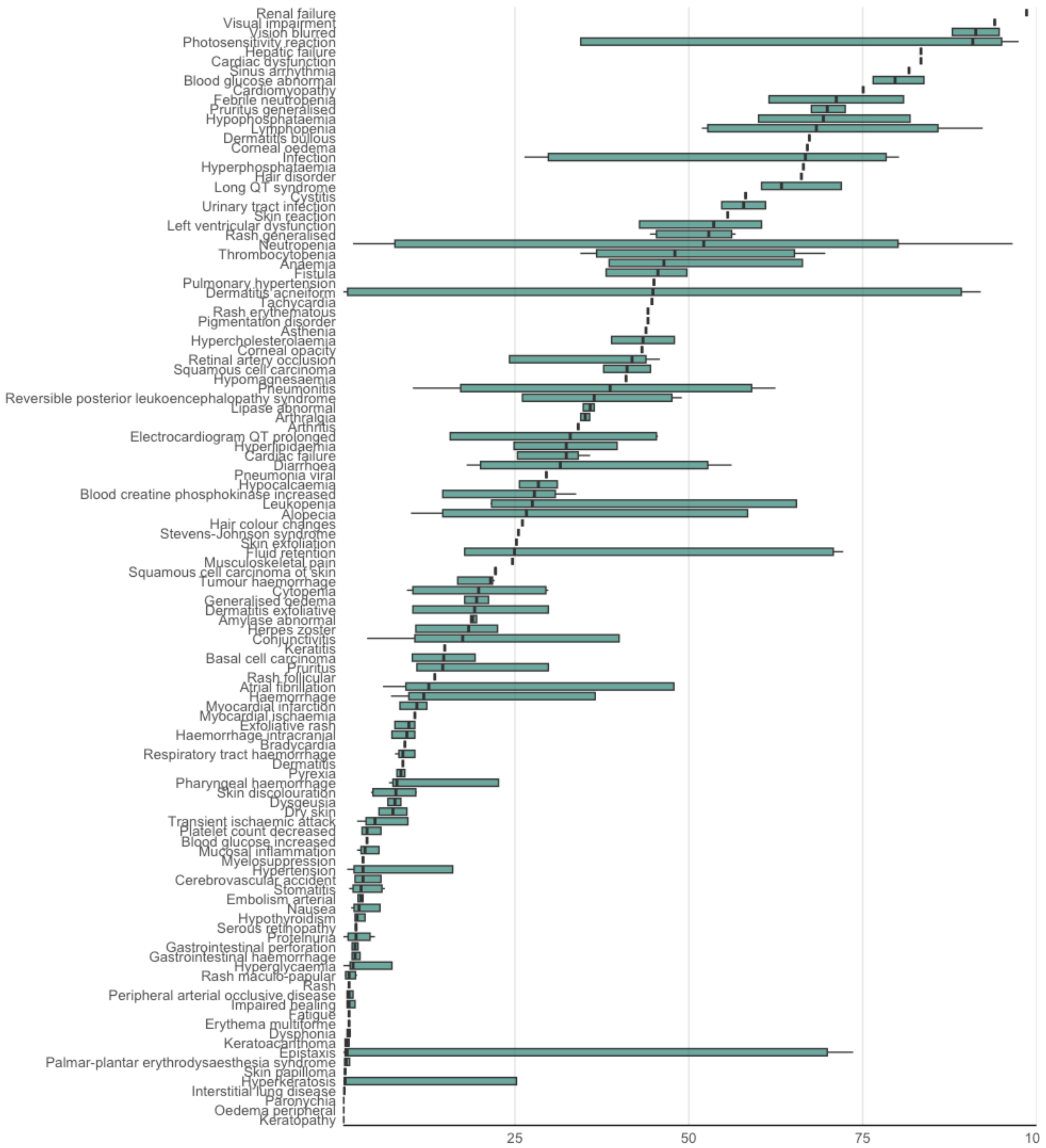


**Figure S4.** Distributions of the percentile ranks of benchmark kinases plotted for each AE at the PT level of the MedDRA hierarchy.

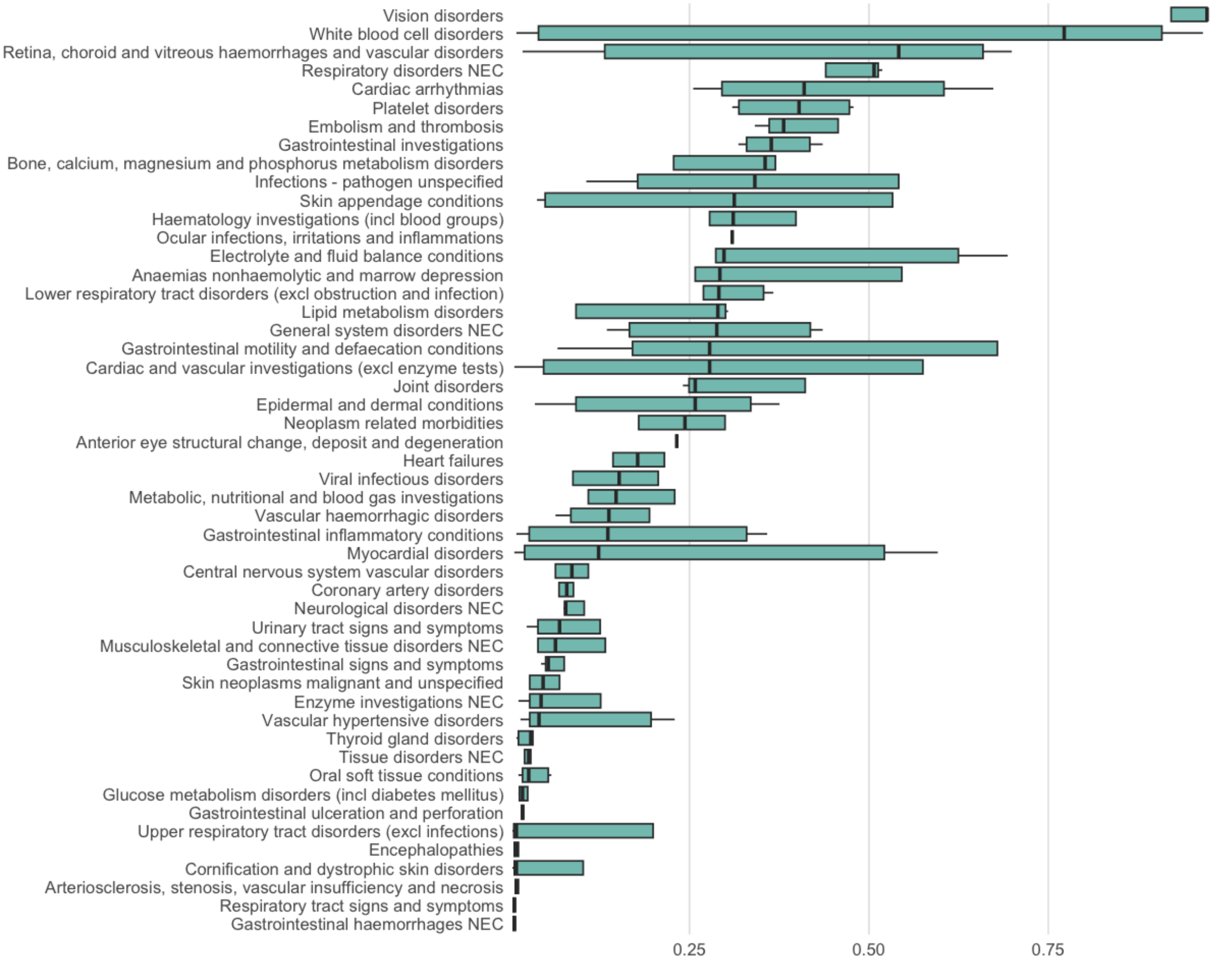


**Figure S5.** Distributions of the percentile ranks of benchmark kinases plotted for each AE at the HLGT level of the MedDRA hierarchy.
